# Relationships of Prescription Opioid Sales to Opiate Treatment Admissions and Overdose Deaths: A Reanalysis of Relationships -- United States 2010-2019

**DOI:** 10.1101/2022.02.23.22271370

**Authors:** Larry Aubry, B. Thomas Carr

## Abstract

**Background:** In 2016, the United States Centers for Disease Control and Prevention (CDC) issued “CDC Guideline for Prescribing Opioids for Chronic Pain”. The guideline used data from 1999 to 2010 that related Opioid Sales to Opiate Treatment Admissions (a measure of drug addiction rates) Opioid Deaths. The CDC’s analysis indicated that opioid prescriptions are directly correlated^1-8^ with drug addiction and overdose mortality outcomes. As a result, prescription opioid sales have been curtailed, making it difficult for sufferers of severe chronic pain to access an effective treatment option. In this paper, the relationships between opioid sales, drug addiction, and opioid deaths are reexamined using data from 2010 to 2019.

**Methods:** Linear regression models were fit to each response separately. Opioid sales (measured as MME per capita) was the independent variable. Total overdose deaths (TOD), any opioid overdose deaths (AOD), prescription opioid overdose deaths (POD) and opiate treatment admissions (OTA) were the dependent, response variables. The models were assessed using three criteria: the statistical significance of the model (Overall P-Value), the quality of the fit (R^2^), and the sign of the slope coefficient.

**Results:** The analyses revealed that the direct correlations (significant, positive slopes) reported by the CDC based on data from 1999 to 2010 no longer exist based on data from 2010 to 2019. The relationships either have reversed (i.e., significant negative slopes) or are non-existent (no significant model).

**Conclusions:** The basis for the CDC guideline no longer holds. The guideline should be updated or annulled based on the current relationships that have existed for a decade. This is especially relevant for people suffering from severe, chronic pain, whose access to prescription opioids has been severely curtailed under the current guideline and to ensure that government resources are directed to an effective approach to reducing overdose deaths.

## BACKGROUND AND RATIONALE

In 2015, the U.S. Department of Health and Human Services (HHS) declared “There is a clear correlation between opioid prescribing rates and overdose death rates in the United States.”^9^ Direct correlations that existed between Total Overdose Deaths (TOD), Any Opioid Deaths (AOD), Prescription Opioid Deaths (POD), Opiate Treatment Admissions (OTA) and prescription opioid sales (POS) from 1999-2010 (see Figure 1) led the CDC to conclude that prescribed opioids are the determinant for overdose deaths and opiate/opioid treatment admissions.^1-8^ Since the 2016 “CDC Guideline for Prescribing Opioids for Chronic Pain” (guideline), cutting prescription opioid sales has been CDC’s, DEA’s, legislative policy makers’, and practitioners’ solution to cut overdose deaths and opiate treatment admissions.^10-12^

**Figure 1:**
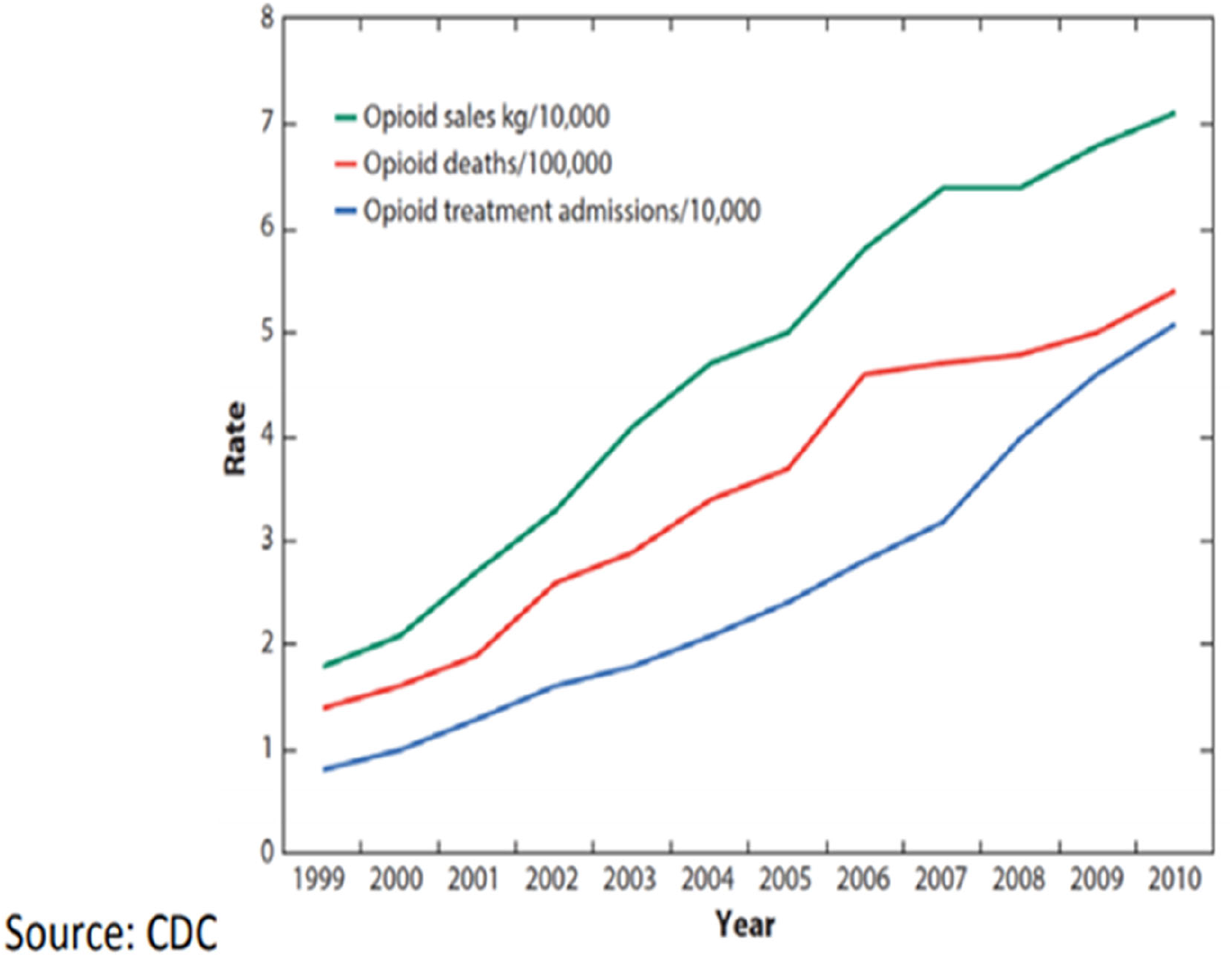
CDC Chart 1999-2010, February 28, 2018, Congressional testimony “Combatting the Opioid Crisis.” made before the Committee on Energy &Commerce, Subcommittee on Health U.S. House of Representatives: “The CDC has shown that a sharp increase in prescriptions for opioids resulted in a corresponding rise in addiction and overdose deaths. This is a CDC graph. The green line represents opioid prescribing, the red line represents opioid deaths, and the blue line represents opioid addiction. The green line went up as opioid prescriptions started to soar, it led to parallel increases in addiction and overdose deaths.”^7^

The impact of the CDC guideline has been systemic. Long term opioid therapy patients are not accepted as new patients by over 40% of primary pain clinics.^12^ Projected cuts to opioid Rx MME per capita will result in 2020 values falling close to levels prescribed in 2000 of 298^13^ while the over 55 population has increased by 40 million. The American Medical Association (AMA) reports “the CDC Guideline has harmed patients”^14^, “72% of pain medicine specialists said that they—or their patients—have been required to reduce the quantity or dose of medication they have prescribed”^14^ as a result of the guideline.

“Almost 18 million Americans are currently taking long-term prescription opioids”^15^ to treat chronic pain. A “Health and Human Services Pain Management Best Practices Inter-Agency Task Force has determined that tens of millions of Americans rely on legal prescription opioids to treat acute or severe chronic pain, including pain arising from cancer as well as terminal or degenerative illnesses. The Food and Drug Administration (FDA) long ago approved opioid medications for these purposes, and doctors throughout the country lawfully prescribe them.”^16^ 92% of physicians and patients believe that opioids reduce pain and 57% report better quality of life.^17^

With drug overdose deaths “growing, inexorably and exponentially, for four decades”^18^ to a record 93,000 in 2020; FY2019 federal funding to combat the opioid crisis increasing to a record $7.6 billion^19^ and “drug overdoses are now costing the United States approximately $1 trillion annually”,^20^ it is critical that public health policy guidance not be based on out-of-date and/or misleading information.

The present paper updates correlations between POS to OTA, POD, AOD and TOD from 2010 to 2019 and shows that the direct relationships that existed from 1999 to 2010 are no longer present -- in fact, in some instances the relationships have reversed. Consequently, we find that continuing the policies of cutting prescription opioid sales to reduce TOD, AOD, POD, and OTA is unfounded.

## METHODS

### DESCRIPTION OF DATA SOURCES

#### Data Limitations

The authors acknowledge that the data reported by CDC and other sources necessarily contains inaccuracies. It is widely understood that it is impossible to ascertain the specific cause of an overdose death in all cases. This may be due, for example, to poly drug use with alcohol, and an inability to reliably determine the source of opioids detected in postmortem blood toxicity screens (e.g., prescription vs. illicit), among other confounding issues.^,21,22^ However, the authors have employed the same data sources that CDC guideline appears to be based upon. As such, the results of analyses presented here are at least as reliable as what the CDC would be able to obtain from their own analyses, if they chose to undertake them. Thus, the following sources have been applied.

#### Drug Overdose Deaths (National); Total Overdose Deaths, Any Opioid Overdose Deaths and Prescription Opioid Overdose Deaths^23^

1999-2019 data accessed from Drugabuse.gov., Published 2021. Accessed January 10, 2021 from https://www.drugabuse.gov/sites/default/files/Overdose_data_1999-2019.xlsx

#### Opioid Overdose Death Crude Rates (U.S. States)^24^

1999-2019 data accessed from CDC, National Center for Health Statistics. Underlying Cause of Death, 1999-2019 were sourced from CDC WONDER Online Database, released in 2020. Data are from the Multiple Cause of Death Files, 1999-2019, as compiled from data provided by 57 vital statistics jurisdictions through the Vital Statistics Cooperative Program. Identified using underlying cause-of-death codes X40–X44, X60–X64, X85, and Y10–Y14. Accessed Feb 7, 2021, 12:01:39 PM from http://wonder.cdc.gov/ucd-icd10.html

#### Opiate/Opioid Treatment Admissions (National)^25^

2006-2008 data accessed from Substance Abuse and Mental Health Services Administration, Center for Behavioral Health Statistics and Quality, “Treatment Episode Data Set (TEDS): 2000-2010”. National Admissions to Substance Abuse Treatment Services. DASIS Series S-61, HHS Publication No. (SMA) 12-4701. Rockville, MD: Substance Abuse and Mental Health Services Administration (samhsa.gov), 2012. p.43. Accessed April 18, 2021 from Treatment Episode Data Set (TEDS) 2000 - 2010 (samhsa.gov)

#### Opiate/Opioid Treatment Admissions (National)^26^

2008-2018 data accessed from Substance Abuse and Mental Health Services Administration, Center for Behavioral Health Statistics and Quality, “Treatment Episode Data Set (TEDS): 2018.”

Admissions to and Discharges from Publicly Funded Substance Use Treatment. Rockville, MD: 2018 TEDS Annual Report. Substance Abuse and Mental Health Services Administration (samhsa.gov), 2020. Table 1.1a. Accessed April 18, 2021 from https://www.samhsa.gov/data/sites/default/files/reports/rpt31097/2018_TEDS/2018_TEDS.html# PSU. 2018 TEDS Annual Report (samhsa.gov)

**Table A1:** Summary of Regression Models Fit in the Paper.

| Dependent Variable | Independent Variable | Years | Related Figure | n | R<sup>2</sup> | P-Value | Slope | 95% LCL | 95%UCL | Interpretation |
| --- | --- | --- | --- | --- | --- | --- | --- | --- | --- | --- |
| MME per Capita | Total Annual Prescription Opioid Sales | 2006-2010 | NA | 5 | 0.94 | 0.006 | 141 | 76 | 206 | Strong Model. Significant, Positive Relationship. |
| Total Overdose Deaths | MME per Capita | 2006-2010 | NA | 5 | 0.97 | 0.002 | 20 | 14 | 26 | Strong Model. Significant, Positive Relationship. |
| Any Opioid Overdose Deaths | MME per Capita | 2006-2010 | NA | 5 | 0.99 | 0.000 | 20 | 17 | 24 | Strong Model. Significant, Positive Relationship. |
| Prescription Opioid Deaths | MME per Capita | 2006-2010 | NA | 5 | 0.97 | 0.002 | 15 | 10 | 20 | Strong Model. Significant, Positive Relationship. |
| Opioid Treatment Admissions/1000 | MME per Capita | 2006-2010 | NA | 5 | 0.91 | 0.011 | 0.60 | 0.26 | 0.94 | Strong Model. Significant, Positive Relationship. |
| Total Overdose Deaths | MME per Capita | 2010-2019 | Figure 3 | 10 | 0.89 | 0.000 | -85 | -109 | -61 | Strong Model. Significant, Negative Relationship. |
| Any Opioid Overdose Deaths | MME per Capita | 2010-2019 | Figure 3 | 10 | 0.92 | 0.000 | -76 | -95 | -57 | Strong Model. Significant, Negative Relationship. |
| Prescription Opioid Deaths | MME per Capita | 2010-2019 | Figure 3 | 10 | 0.03 | 0.615 | -1.4 | -7.4 | 4.7 | No Model. Nonsignificant Relationship. |
| Opioid Treatment Admissions/1000 | MME per Capita | 2010-2018 | Figure 3 | 9 | 0.89 | 0.000 | -0.64 | -0.84 | -0.43 | Strong Model. Significant, Negative Relationship. |

#### Opioid Prescribing; MME per Capita (National)^27^

2006-2013 data accessed from CDC,” Annual Surveillance Report of Drug-Related Risks and Outcomes — United States Surveillance Special Report”.2019 CDC, U.S. Department of Health and Human Services. Published November 1, 2019. p.115. Accessed January 10, 2021 from https://www.cdc.gov/drugoverdose/pdf/pubs/2019-cdc-drug-surveillance-report.pdf

#### Opioid Prescribing; MME per Capita (National)^28^

2014-2018 data accessed from Statista,” Annual morphine milligram equivalents (MME) dispensed per capita in the U.S. from 2014 to 2018”, MME per capita U.S. 2014-2018. Statista. May 28, 2021. Accessed July 8, 2021 from https://www.statista.com/statistics/753284/number-of-mme-dispensed-per-capita-in-us/.

#### Opioid Prescribing; MME per Capita (National)^29^

2019 data accessed from The IQVIA Institute, “Prescription Opioid Trends in the United States”, Institute Report, Dec 16, 2020. p.4. Accessed January 10, 2021 from https://www.iqvia.com/insights/the-iqvia-institute/reports/prescription-opioid-trends-in-the-united-states.

#### Opioid Prescribing; Opioid Dispensing Rates per 100 (U.S. States)^30^

2006-2019 data accessed from CDC, “U.S. Opioid Dispensing Rate Maps, Drug Overdose”, CDC Injury Center. Accessed February 9, 2021 from https://www.cdc.gov/drugoverdose/rxrate-maps/index.html

#### Opioid Sales kg/10,000 (National)

For the period from 2006 through 2018/2019, these data were not known to be publicly available. We instead examined Opioid Prescribing by separately computing MME per Capita.

### STATISTICAL METHODOLOGY

#### Objective 1: Evaluate MME per Capita as a legitimate alternative measure of Annual Prescription Opioid Sales

The CDC used Annual Prescription Opioid Sales to support their current guideline (Figure 1). Data on Annual Prescription Opioid Sales are not readily available since 2010. However, MME per Capita data are available from 2006 to 2019 and offer a reasonable surrogate. Annual Sales data from the CDC chart were visually extracted and correlated with MME per Capita data, using simple linear regression analysis. The goal of the analysis was to evaluate MME per Capita as a legitimate alternative measure of Annual Prescription Opioid Sales.

#### Objective 2: Assess the strength and nature of the relationships between total overdose deaths (TOD), any opioid overdose deaths (AOD), prescription opioid overdose deaths (POD) and opiate treatment admissions (OTA) and Opioid Sales/MME per Capita

Consistent with the methods used by the CDC, simple linear regression models were fit to the data. Separate models were fit to each of the four dependent variables (TOD, AOD, POD, and OTA) using Annual Opioid Sales (i.e., MME per Capita) as the independent variable. Two models were fit to each dependent variable. One model covered the years presented in the original CDC chart (for which MME per Capita data were available) (2006-2010) and the second model covered the years since the published CDC chart (2010-2019).

For both objectives, the strength and nature of relationships in all the regression models were assessed using three criteria:

1. significance of the regression model (overall P-Value),
2. the quality of the model’s fit (R^2^), and
3. the sign of the linear slope coefficient (+ or -).

All models were fit using PROC REG from SAS/STAT software Version 9.4.

## RESULTS AND DISCUSSION

Data from CDC’s original chart was reconstructed using graphical analysis. The reconstructed Annual Prescription Opioid Sales values from the original CDC chart are highly correlated with publicly available MME per Capita values (R^2^ = 94%), which are available for more recent years than the sales data used by the CDC. MME per Capita is thus used in place of Annual Prescription Opioid Sales for all subsequent analyses.

For the years covered in the CDC’s original chart (for which MME per Capita data are available, i.e., 2006-2010), the CDC’s claim of positive/direct relationships between TOD, AOD, POD, and OTA and Annual Prescription Opioid Sales (i.e., MME per Capita) were validated (91% < R^2^ < 97%), with statistically significant, positive slopes.

For more recent years (i.e., 2010-2019), however, the CDC’s assertion of continued direct relationships is not valid. The relationships between TOD, AOD, POD, and OTA and Annual Prescription Opioid Sales (i.e., MME per Capita) are either non-existent or significantly negative/inverse (Figures 2 and 3).

**Figure 2:**
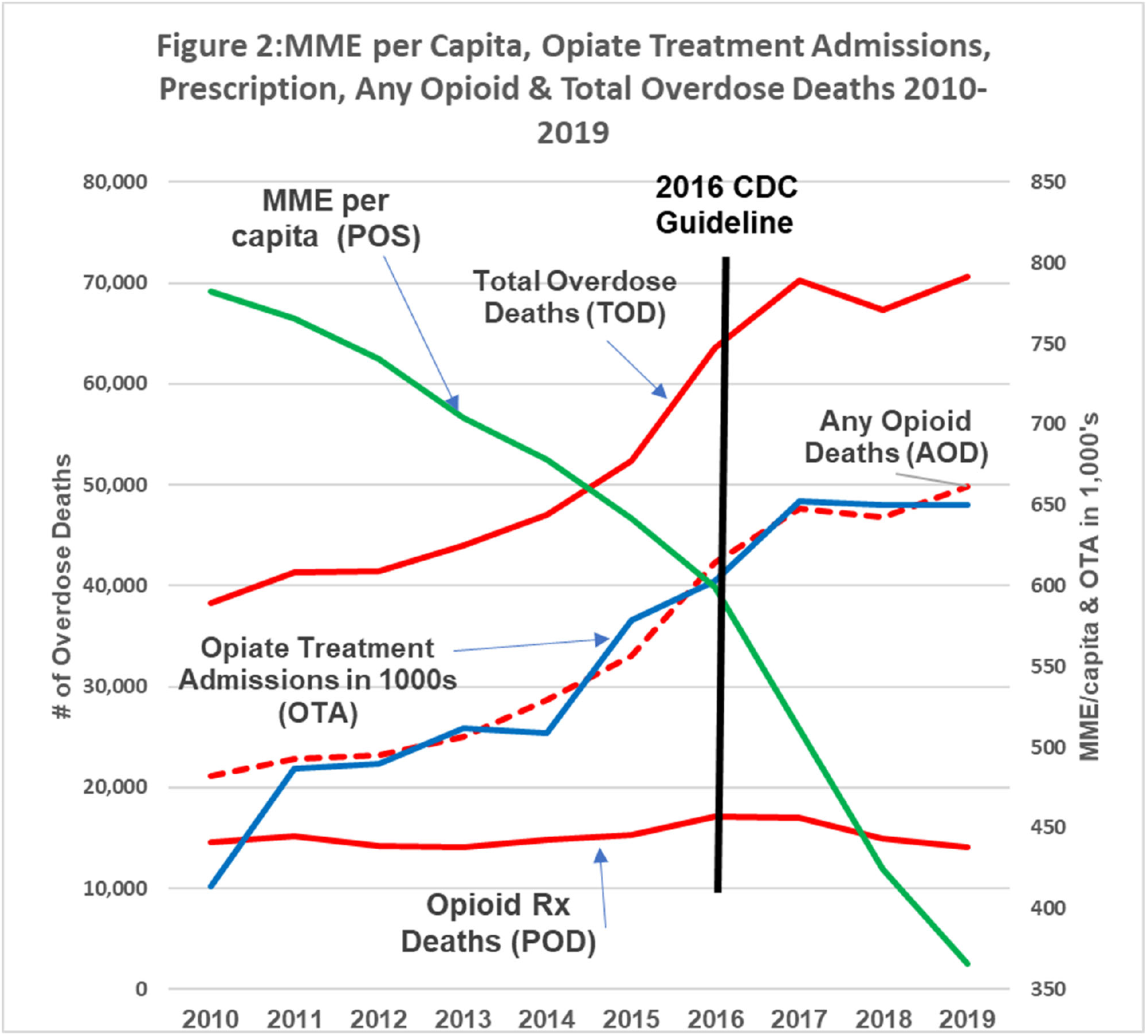
2010-2019 Update: The green line represents opioid prescribing (POS, MME/capita); the red lines are opioid deaths (POD, AOD &TOD); the blue line represents opioid addiction (OTA). Over the past decade, as the green line (prescription opioids) declined by +50%, prescription opioid deaths remained flat while opioid addiction, any opioid and total overdose deaths continued increasing “exponentially”^8^.

**Figure 3:**
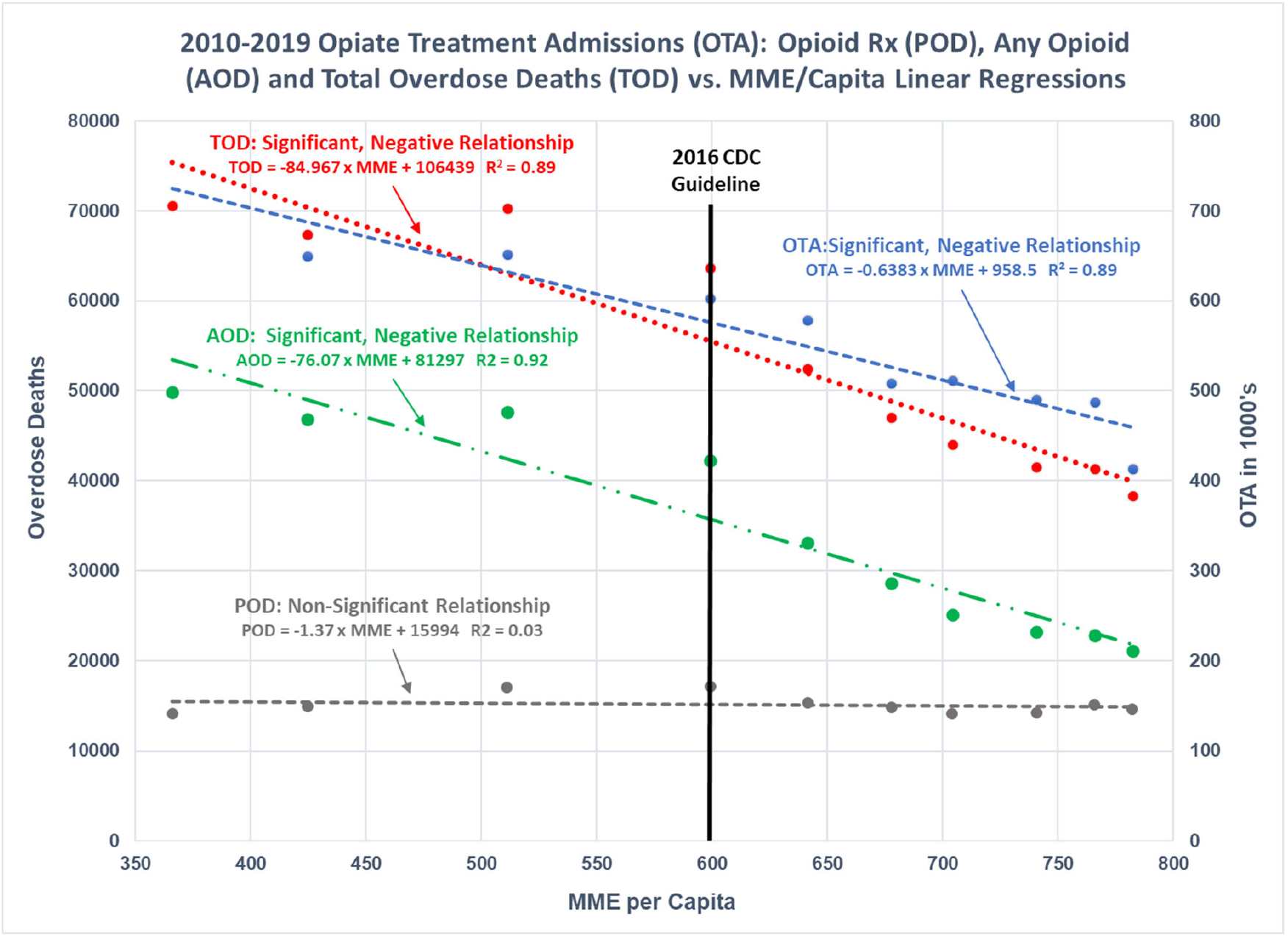
2010-2019 Regression Models: Illustrates the regression of OTA, POD, AOD and TOD as functions of POS. Significant, negative relationships were found for OTA, AOD, and TOD. No significant relationship exists between POD and POS.

National trends since 2010 are paralleled in a strong majority of states. Between 2010 and 2019 inclusive, there was a statistically significant negative correlation (95% confidence level) between AOD and Annual Prescription Opioid Sales in 40 states, with significant positive correlations occurring in only 2 states. Eight (8) states did not exhibit a significant (95% confidence level) relationship between overdose deaths and prescription opioid sales during the 2010-2019 time period.

Results for all regression models are presented in Table 1.

## CONCLUSIONS

Starting in 2010 opioid MME per Capita (POS) does not have a “clear correlation”^9^ or move “in parallel”^3^ or “in lockstep”^2^ with OTA, POD, AOD or TOD. The relationship changed from direct to inverse.

From a peak in 2010 to 2019, POS decreased by over 50%. If POS were the root cause, correlated to and resulted in “parallel” or “lockstep” movements in POD, AOD, OTA, and TOD, then the result should have been an equivalent percentage reduction in these metrics. From 2010 to 2019, POD should have been 6,800 in 2019 instead of ending the decade flat at 14,139; AOD should have been 9,900 instead of increasing by 2.5x to 49,860; OTA should have been less than 210,000 in 2018 instead of increasing by 1.6x to 650,212 and TOD should be 17,900 instead of doubling to 70,630.

Data from the National Health Interview Survey (NHIS) demonstrate that between 2011 to 2016, while MME per capita declined by 21.8%, the amount of pain reported by Americans significantly increased. Incidence of high impact chronic pain increased by 84.9%, from 4.8 percent (10.6 million people, 1 in 20)^31^ to 8.0% of U.S. adults (19.6 million, 1 in 12.5)^32^. Incidence of patients afflicted with chronic pain increased by 67.2% from 13.6 percent (29.9 million, 1 in 7.4)^31^ to 20.4% (50.0 million, 1 in 5)^32^ of the U.S. adult population.

In light of the evidence presented in this paper, it should be recognized that the basis for the 2016 guideline is no longer valid. The guideline and associated content should be updated or annulled based on the current relationships that have existed for a decade, to eliminate the unnecessary suffering of individuals with chronic pain and to avoid the wasteful and ineffective spending patterns the government is executing based on the out-of-date guideline.

## Data Availability

All data produced in the present study are available upon reasonable request to the authors

